# Post COVID-19 conditions in Children and Adolescents at 3 months following a Delta outbreak in Australia: a cohort study

**DOI:** 10.1101/2023.03.14.23287239

**Authors:** Philip N Britton, Rebecca Burrell, Emily Chapman, Julia Boyle, Shirley Alexander, Yvonne Belessis, Jacqueline Dalby-Payne, Katherine Knight, Christine Lau, Brendan McMullan, Bronwyn Milne, Marilyn Paull, Jonathan Nguyen, Hiran Selvadurai, Russell Dale, Andrew Baillie

## Abstract

**Background:** Long COVID remains incompletely understood in children and adolescents with scant Australian data available. We aimed to assess the impacts of the 2021 Delta variant of SARS-CoV-2 outbreak on symptoms and functioning 12 weeks post-acute infection in a cohort of children and adolescents.

**Methods:** The parents (or next of kin) of 11864 children and adolescents from a population catchment who had mandatory contact with Sydney Children’s Hospital Network facilities during acute SARS-CoV-2 infection (confirmed by PCR) were contacted by email or text message.

**Findings:** 1731 (17.7%) responded to an online survey assessing symptoms, functional impairment. 203 of the responders (11.7%) gave answers that were consistent with continued symptoms and/or functional impairment and were flagged for clinical review. Of the 169 subsequently clinically reviewed, many had already recovered (n=63, 37.3%) or had a pre-existing condition exacerbated by COVID-19 (18, 10.7%); 64 (37.9%) were diagnosed with a Post COVID Condition (PCC). Of these, a minority we considered to have features compatible with the United Kingdom consensus cases definition for Long COVID (n=21).

**Interpretation:** During an outbreak of the Delta variant of SARS-CoV-2 an online questionnaire with clinical review follow-up provided evidence that a majority of children with COVID-19 had complete recovery at 12 weeks post infection, but those with persisting symptoms demonstrated a wide spectrum of severity and phenotype that comprises a likely significant burden that warrants attention for individuals and at a population level.

**Funding:** New South Wales Health COVID-19 Emergency Response Priority Research Funding.

## Background

Post-COVID-19 conditions including Long COVID remain poorly understood in children and adolescents. Systematic reviews and guidelines have highlighted potential multi-system involvement beyond the period of acute SARS-CoV-2 infection, with more than 200 symptoms reported.^1,2,3^ However, heterogeneity and biases across various studies have complicated interpretations of prevalence, clinical spectrum, trajectory, and burden of functional and socio-economic impacts.^1,2,4^ In adults the impact of Long COVID is likely to be substantial, with more than one third of hospitalised people in the UK reporting reduced quality of life following acute COVID-19 in an unvaccinated cohort^2^, nearly half working reduced hours, and one fifth still unable to work several months after the infection.^5^

The pathogenesis of Long COVID is currently considered to be related to all, or any of, the following: residual organ damage from the acute illness, persistence of viral antigens, prolonged systemic inflammation and vascular injury, an immune response with inflammatory cytokine production associated with endothelial dysfunction and a procoagulant state.^1^ In adults, recovery from COVID-19 is influenced by comorbid conditions, psychological impacts of the acute illness, social isolation and/or life-style changes enacted during the pandemic.^6^ In the absence of specific diagnostic tests, Long COVID in adults has been recognised to occur in symptom clusters albeit variably defined in different studies.^2,7,8^

Pre-morbid factors, such as obesity, diabetes and ethnicity (albeit confounded by socio-economic inequities), are strongly associated with severe acute COVID-19,^9^ but predictors of Long COVID have not been well defined.^2^ Other potentially important risk factors include body mass index (BMI), depression, and childhood hardship.^2,5^

Long COVID is likely less frequent in children and adolescents compared with adults, and there is concern that the social and economic – sometimes called indirect effects - of the ‘Pandemic’ on the health and wellbeing of children and adolescents has been substantial and need to be accounted for in determining the burden of long COVID.^10,11^ Long COVID shows some clinical similarity to other post-viral fatigue syndromes which are currently being used to guide management whilst further information emerges regarding pathogenesis and treatment.^12^ Increasingly, Long COVID is being considered a sub-set of those persons with post-acute sequelae of COVID-19 (PASC) that make up a heterogenous group of post-COVID conditions (PCC). Recent publication of a consensus case definition for Long COVID in children (Box 1) is well aligned with the WHO case definition for adults, and will likely inform future research studies.^13^

There have been scant data published on PCC in children in Australia.^14^ Here, we aimed to determine the prevalence and describe the clinical spectrum of PCC amongst children and adolescents following the 2021 Delta variant of SARS-CoV-2 outbreak in New South Wales (NSW), Australia. NSW is Australia’s most populous state and had the largest Delta variant of concern (VoC) outbreak.

### Box 1

Consensus Definition of Long COVID in Children^12^

A condition in which a child or young person has symptoms (at least one of which is a physical symptom) that:

- Have continued or developed after a diagnosis of COVID-19 (confirmed with one or more positive COVID tests)
- Impact their physical, mental or social wellbeing
- Are interfering with some aspect of daily living (e.gg, school, work, home or relationships) and
- persist for a minimum duration of 12 weeks after initial testing for COVID-19 (even if symptoms have waxed and waned over that period)

## Methods

### Participants

During the Delta VoC outbreak in 2021, the Sydney Children’s Hospitals Network (SCHN) supported the outpatient management of around two thirds of all notified cases of SARS-CoV-2 infection in the Australian state of New South Wales through its ambulatory care service, virtualKIDS-COVID outpatient response team (VK-CORT). Incidence of hospitalisation and acute clinical spectrum of disease in children has been previously described.^15^ A great majority of children had mild or uncomplicated disease and were cared for in the community. ^15^

During the outbreak, SCHN clinicians expressed a need to determine the medium-term outcome of these cases given emerging evidence at the time regarding Long COVID in children. However, the scale of the outbreak resulted in considerable capacity constraints around clinical outcome assessment. We devised a digital solution to facilitate outcome assessment using REDCap^16^ to send a questionnaire to as many cases as possible (Figure 1).

**Figure 1:**
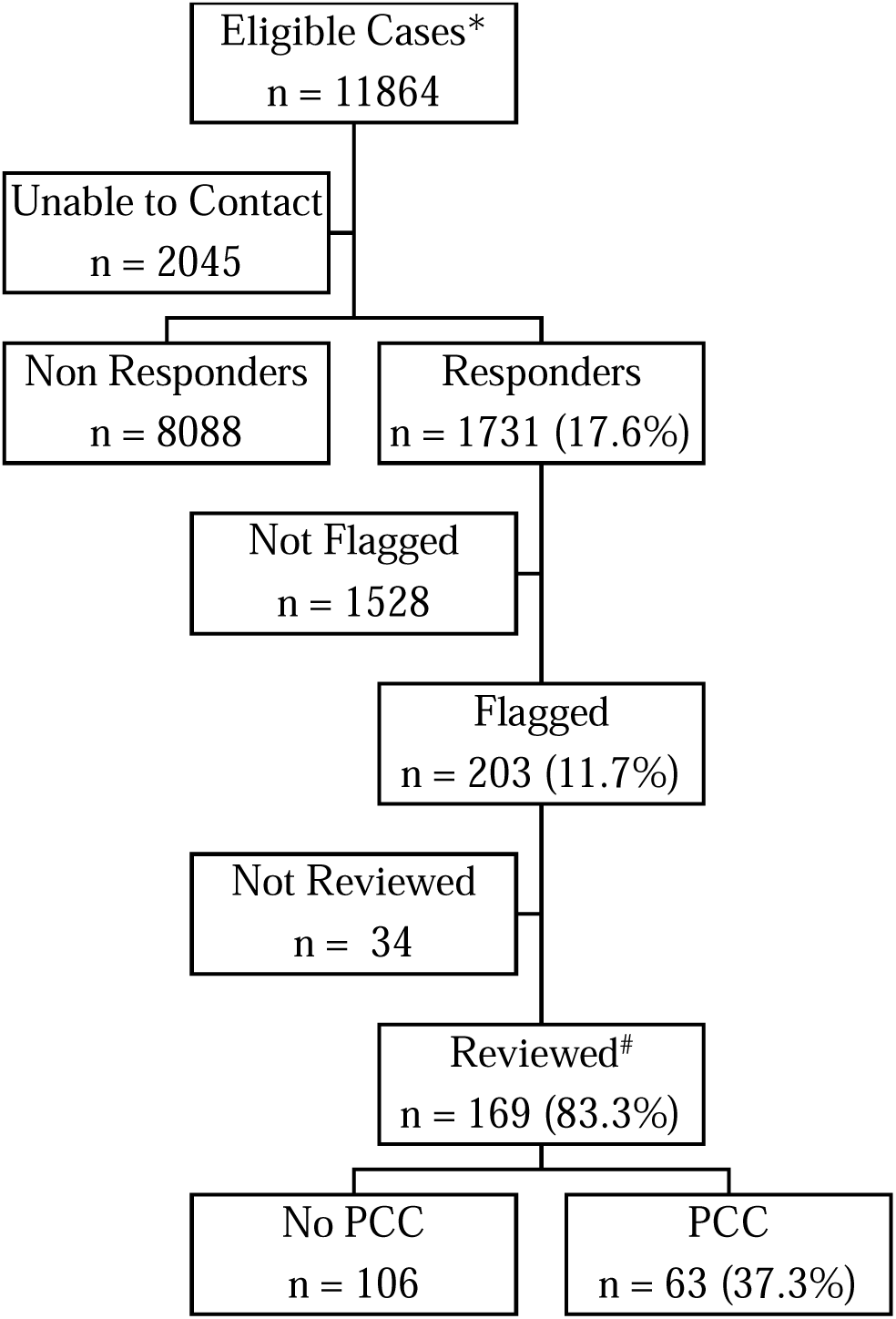
STROBE^9^ style participant flowchart of eligible children and young people invited to participate and respondents. *Eligible cases were identified in Western, South Western, and South Eastern local health districts via confirmed SARS-CoV-2 infection and referral to the SCHN virtualKIDS□CORT (as previously described).^15^ A total of 38 cases of incorrect referrals were identified and removed (reasons included aged over 20 years (n=20), false positive result (n=16), incorrect patient matching (n=2)). A further two children were removed due to social admission to virtualKIDS-CORT and remained test negative. Of responders 63 (3.6%) children responded for themselves; 1691 (96.1%) of responses were from a parent, guardian, or other family member (mother 1364, 81.1%; father 270, 16.1%; grandparent 21, 1.2; sibling 12, 0.7%; aunt, uncle or other 14 0.8%); with 5 (0.3%) interviews with one of the researchers.

### Outcome measures

We developed a combined 15-minute online questionnaire based on: the International Severe Acute Respiratory and emerging Infection Consortium (ISARIC) Global Tier 1 Ongoing COVID-19 Follow Up Survey including the ISARIC symptom list; the age-specific EuroQOL questionnaire (EQ5D-Y) for health-related quality of life^17^; selected questions from the World Health Organisation disability assessment score (WHO-DAS)^18^ for functional impairment; and, the Kessler K6^19^ as a screen for mental health or Parent and/or Adolescent Strengths and Difficulties Questionnaire (SDQ) ^20^. Importantly, we began the questionnaire with four screening questions to ensure assessment of outcome even where the full questionnaire was not completed.

The questionnaire was distributed to a panel of specialists - the ‘Long COVID clinical panel’ comprising senior clinicians from general paediatrics, infectious diseases, respiratory medicine, adolescent medicine and psychological medicine - within SCHN for review and comment and pre-determined flags for clinical concern were set (Supplementary methods). These were then set as pre-determined parameters for digital ‘flagging’ of responses in REDCap that would require clinical review.

### Procedures

The REDCap (Sydney Local Health District (SLHD) server) based questionnaire was automatically sent to families 12 weeks following registration in VK-CORT following bulk upload of contact details (email, mobile phone) onto the SLHD server. The questionnaire was sent to all SCHN VK-CORT patients who tested positive for SARS-CoV-2 between July-November 2021 using a listed parent/carer email address (from 15/11/2021-10/02/2022) and/or mobile number (from 22/12/2021-18/02/2022).

Early in the follow-up process, we recognized that more than half of the flagged responses were for raised SDQ scores only. Following direct contact with the first 20 of such cases by a Child and Adolescent Psychiatrist, it was determined that the SDQ identified pre-existing emotional and behavioral issues rather than screening for new symptoms of concern that may be associated with Long COVID.

Of the cases with responses showing flags for clinical concern beyond SDQ responses, a parent/carer was contacted by a paediatric clinician for a telehealth review of progress and clinical validation of any concerns. These clinical reviews were undertaken by members of a multi-specialty ‘Long COVID clinical panel’ and independently documented in clinical notes. Clinical documentation of these assessments was reviewed by a paediatric infectious diseases physician (PB) and paediatric clinical nurse consultant (EC) and any issues identified were pragmatically categorized into five clinical groups: Mental health issues, Pre-existing condition (exacerbated by COVID-19), Single organ symptoms/dysfunction, Persistent symptoms (multi-organ) and Post-viral fatigue. If a parent/carer reported that the symptoms they recorded in the survey had resolved, the child was categorized as ‘recovered’.

We considered two clinical sub-groups – post-viral fatigue and persistent symptoms (multiorgan) - to be potentially consistent with the UK consensus case definition for ‘Long COVID’ if a reviewing clinician could validate the persistence of symptoms and there was a definite documented impact of the symptoms on function (inability to participate in usual activities and/or non-attendance at school or early childhood education centre, excluding that because of public health orders).

Duplicate questionnaire responses (n=5) were identified and manually checked for discrepancies (PB, RB). Responses were retained for analyses where one questionnaire was more complete (n=4). In the instance both responses were completed with minor discrepancies, the response most proximal to illness was retained for analyses.

REDCap questionnaire data, medical record data, and clinical review data were merged using medical record number. Duplicate cases, identified via MRN, name, and date of birth, were removed prior to merging. Descriptive analyses were completed using IBM SPSS Statistics (Version 25) and Microsoft Excel (2013). We report prevalence of self-reported non-recovery across multiple variables, clinician assessed clinical spectrum amongst those with self-reported non-recovery. We calculated relative risks of non-recovery at clinician review by demographic and clinical variables available during acute illness.

The study received ethical approval from the Sydney Local health District (Royal Prince Alfred Hospital Zone) Human Research Ethics Committee (HREC ID: 2021/ETH11819 & X21-0370).

## Results

Of 11,864 eligible children and families, 9765 had contact details (mobile phone number or email address) extractable from electronic medical records (2099 had blank or nonsense records). Of these, 1825 (18.7%) opened the URL to the REDCap questionnaire and 1731 (17.7%) answered one or more of the four screening questions (Figure 1). We compared those eligible cases who responded with non-responders. The epidemiologic curve of responders and non-responders were broadly overlapping (Supplementary Figure) and there was also broad similarity between the groups in terms of demographic variables and acute disease severity with some exceptions (Supplementary Table 1). The responder cohort showed a higher prevalence of fever and cough at assessment of acute infection, was slightly younger and was less likely to have a postcode of residence from more socioeconomically disadvantged areas ^21^.

Of the responders (n=1731), 443 showed responses that flagged for concern, but upon review around half (240/443) of these were for elevated scores on the SDQ only. The remaining 203 cases (13.7%) of responses were considered as flagged for or self-identified as a possible post-COVID condition, see Figure 1. The responses for pre-determined ‘flagging’ questions from the questionnaire are shown in Table 1. Persistence of COVID-19 associated symptoms was reported in 6.1% (106/1731) of participants, non-recovery to baseline health in 6.0% (104/1728), inability to return to usual (baseline) activities in 3.0% (52/1730) and a need for additional help to recover from COVID-19 in 1.7% (30/1730) of responses. Of those who reported persistent symptoms and went on to complete the full ISARIC symptom check list (n=64), a wide spectrum of symptoms were reported and their frequencies by system are shown in Figure 2, with individual symptom frequencies in Supplementary Figure 2 and Supplementary Table 2. Age-related differences were evident when comparing children above and below an age cut-off at 11 years. All symptom groups were more frequent in older children, with sensory symptoms (loss of taste or smell) and neurological symptoms significantly more frequent (Figure 2)

**Table 1:** Key results for pre-determined ‘flagging’ questions from questionnaire responses.

| Key question (response result) | n/N | % (95% CI) |
| --- | --- | --- |
| *Screening Q* |  |  |
| Is [your child] interacting as per their usual baseline (i.e. before COVID)? (NO) | 104/1728 | 6.0 (4.9 - 7.3) |
| Does [your child] have ongoing symptoms that concern you related to their COVID-19 illness? (YES) | 106/1731 | 6.1 (5 - 7.4) |
| Are [your child’s] COVID-19 related symptoms stopping them from doing their usual activities? (YES) | 52/1730 | 3.0 (2.2 - 3.9) |
| Do you think you need additional help for [your child] to recover from COVID-19? | 30/1730 | 1.7 (1.2 - 2.5) |
| Any screening Q positive | 129/1731 | 7.5 (6.2 - 8.9) |
| *Additional ‘flagging questions’* |  |  |
| ISARIC How much do you agree with the statement, “[My child] has fully recovered from COVID-19”?<br>(0-10 scale with 0=strongly disagree; result <2) | 9/1610 | 0.6 (0.3 - 1.1) |
| Pediatric Dyspnea Scale How much difficulty has [your child] had breathing in the last 7 days? (>=‘some’) | 30/1615 | 1.9 (1.3 - 2.7) |
| WHO DAS How do you rate [your child’s] overall health in the past 30 days? (<=bad) | 8/1576 | 0.5 (0.2 - 1) |
| WHO DAS Q6 Days absent from school/ preschool/ daycare in past 30 (>9) | 33/1380 | 2.4 (1.6 - 3.4) |
| Has [your child] been readmitted to hospital after first acute covid-19 illness | 15/1555 | 1 (0.5 - 1.6) |
| ISARIC symptom list - Greater than 3 symptoms | 28/64 | 43.8 (29.1 - 63.2) |
| EQ-5D-Y Q4 Having Pain or Discomfort (a Lot) | 2/657 | 0.3 (0 - 1.1) |
| Age 12+ K6 Q4 About how often has [your child] felt hopeless (most of the time or all the time) | 6/408 | 1.5 (0.5 - 3.2) |
| Age 12+ K6 total score greater than 18 | 11/410 | 2.7 (1.3 - 4.8) |
| Age 4-12 SDQ Q26 Difficulties in emotions, concentration, behaviour or being able to get on with other people (severe) | 10/687 | 1.5 (0.7 - 2.7) |
| Age 4-12 SDQ flag* | 301/687 | 43.8 (39 - 49.1) |
| Any additional flagging q excluding SDQ flag | 74/1731 | 4.3 (3.4-5.3) |
| Total flagged for clinical review (excluding SDQ flag) | 203/1731 | 11.7 (10.2-13.3) |

Abbreviations: ISARIC, International Severe Acute Respiratory and emerging Infection Consortium; WHO-DAS World Health Organisation Disability Assessment Schedule, K6 Kessler 6 item distress scale, SDQ = Strengths and Difficulties Questionnaire.
\* Strengths and Difficulties Questionnaire flag included any of, SDQ total difficulties scale or subscales in "borderline" or "abnormal" range from original 2-band categorisation.

**Figure 2:**
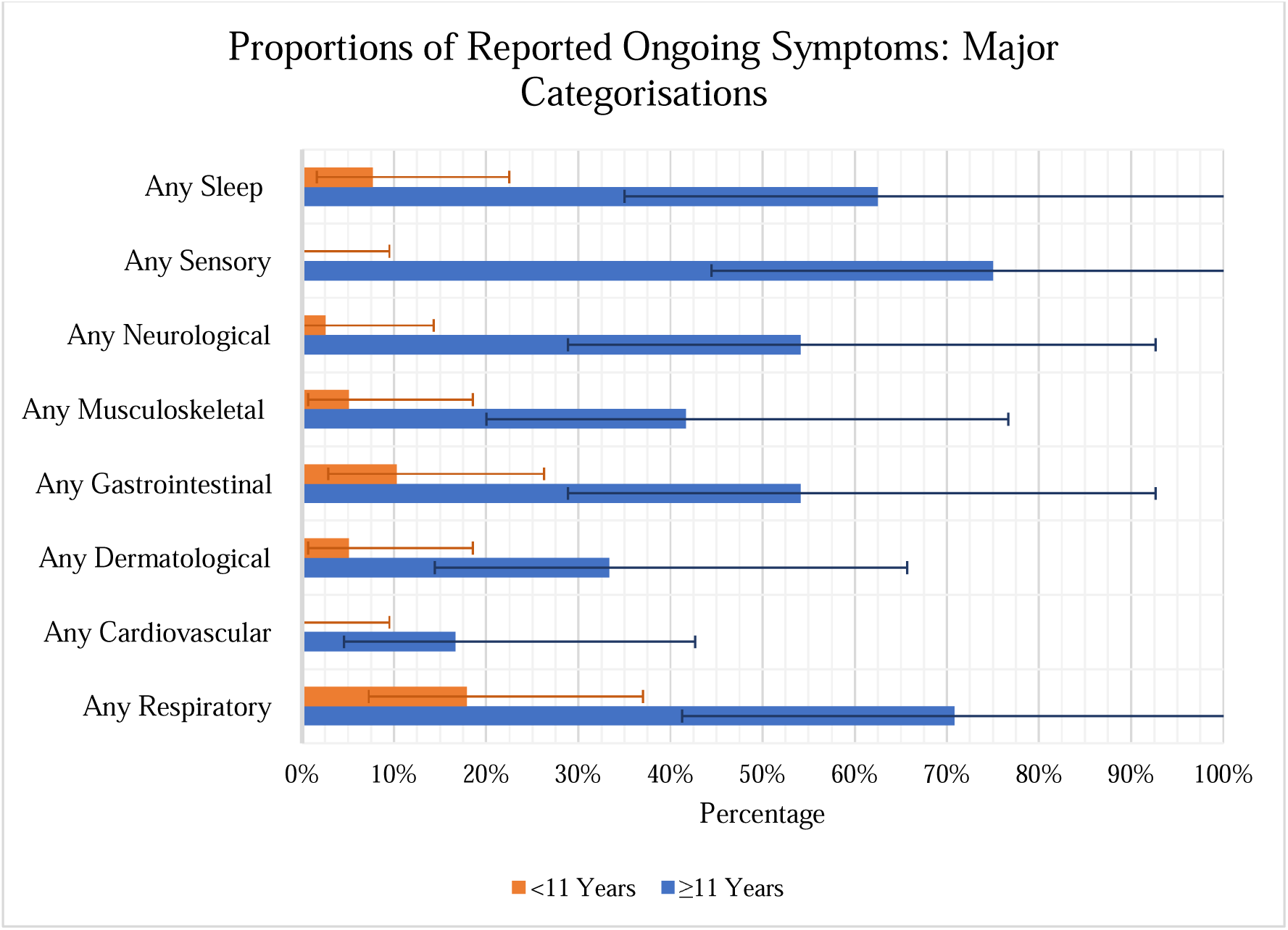
Bar graph of self-reported persistent symptoms ((percentage and 95% confidence intervals) using ISARIC symptom questionnaire symptom list stratified by age; younger children aged <11y, older children aged ≥11y (n=64).

Measures of the overall effect of disease showed 0.5% (8/1576) of responses rating the child’s health as ‘bad’ or worse on the WHO-DAS, 2.4% (33/1380) of responses indicating the child had missed 10 or more days of school/preschool or daycare in the preceding 30 days, and 5.1 % (79/1559) of responses indicating they had ‘visited a doctor/health centre because of COVID-19 health consequences’. In terms of the self-reported “Total Health Score” (maximum score possible = 100) the mean score from responders was 90.7 (IQR 89; 100) prior to acute COVID-19 and 87.8 (IQR 82; 100) on the day of questionnaire completion (Supplementary Table 3).

### Clinical review

Of the 203 responses showing flags for clinical concern, 169 (83%) were able to be contacted by a paediatric clinician for telehealth review (36 unable to be contacted). Of these, 156/203 (77%) had sufficient clinical documentation of a paediatric assessment to support pragmatic clinical case categorization (Figure 1 and Table 2). 63 children and adolescents showed complete recovery at clinical review despite having prior self-reported non-recovery; with a median time from questionnaire completion to clinical review being 23.2 days (IQR 15.4-47.4).We identified 63 children with evidence of a post-COVID condition. Amongst these were children with single-organ persistent symptoms, most prominently upper or lower respiratory tract symptoms, multi-organ persistent symptoms or predominantly fatigue. There were 21 children (1.2% of responder cohort, 95% CI 0.8-1.8) with persistent symptoms of more than one type and confirmed to be impacting on daily function (most often ability to attend school regularly) and so considered compatible with the UK consensus definition of Long COVID.

**Table 2:**
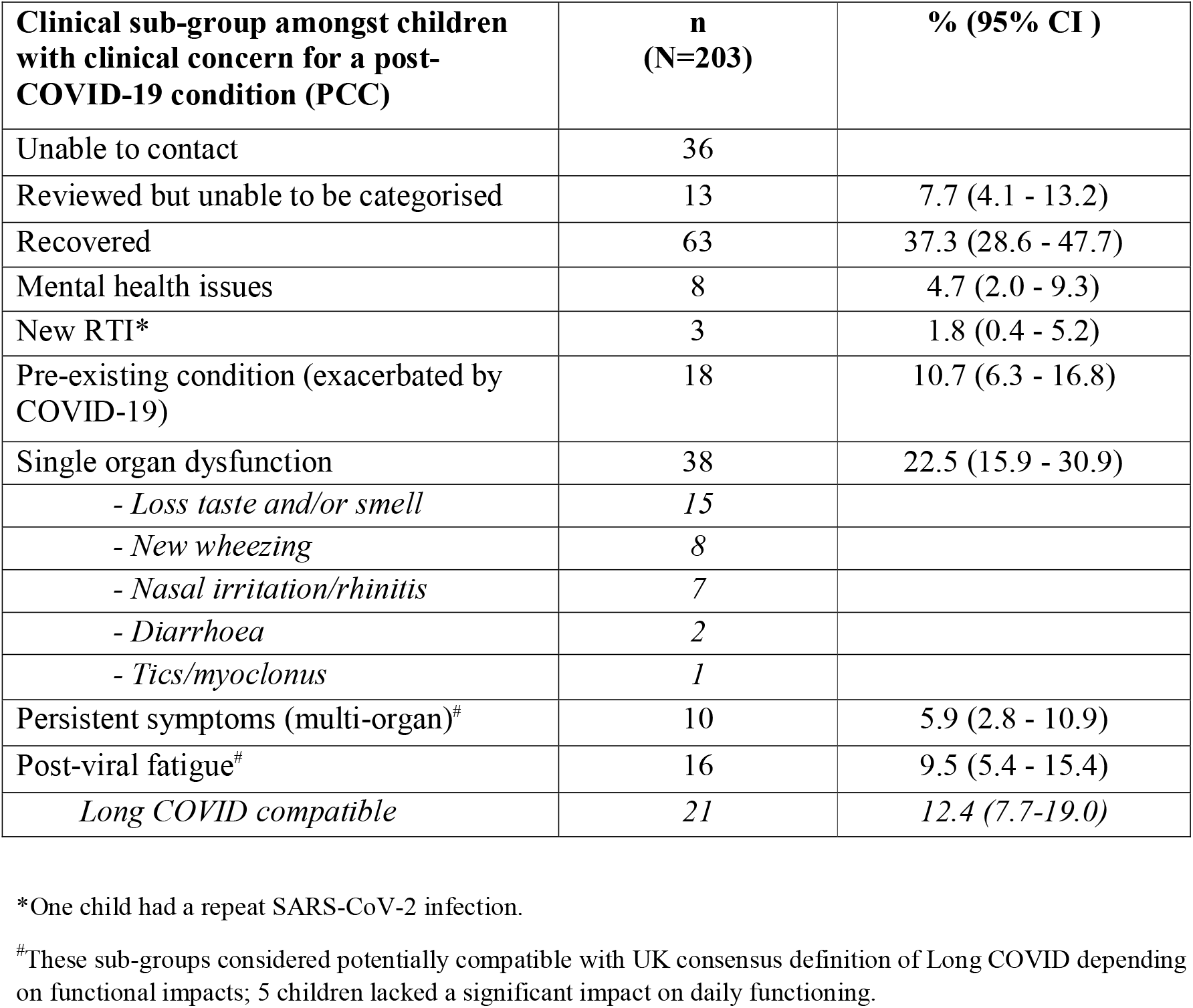
Clinical review with pragmatic clinical categorisation of the 203 children flagged from questionnaire responses.

We compared demographics and acute illness variables between responders who self-reported recovery, and responders with self-reported concern for non-recovery stratified by the outcome of clinical review (Table 3). Those children with clinician confirmed PCC were more likely to be older (relative risk 4.0 (RR), 95% CI 2.0-7.8; 12-15 years compared with <5 years) and more likely to have medical co-morbidity (RR 2.3, 95% CI 1.4-3.7). Children with PCC were also more frequently female and considered more unwell/higher risk acutely than those who self-reported recovery although this was not statistically significant.

**Table 3:** Demographics, symptoms, level of impairment amongst responder groups both flagged for clinical review and not.

|  | **Responders Not Flagged\* (N=1278)** | **Flagged (Self-identified) Not Reviewed (N=34)** | **Flagged (Self-identified) Reviewed but no PCC (N=106)** | **Flagged (Self-identified) Reviewed and confirmed PCC (N=63)** | **RR^ (95%CI)** |
| --- | --- | --- | --- | --- | --- |
| **Gender female** | 651 (50.9) | 14 (41.2) | 35 (38.0) | 37 (58.7) | 1.35 (0.83-2.2) |
| **Age group** |  |  |  |  |  |
| *under 5 years* | 491 (38.4) | 9 (26.5) | 34 (32.1) | 11 (17.5) | Ref |
| *5 to 11 years* | 475 (37.2) | 15 (44.1) | 48 (45.3) | 22 (34.9) | 2.0 (0.99-4.1) |
| *12 to 15 years* | 295 (23.1) | 10 (29.4) | 24 (22.6) | 28 (44.4) | 4.0 (2.0-7.8) |
| *16 or older* | 6 (0.5) | 0 | 0 | 1 (1.6) | 6.5 (0.96-44) |
| *Missing* | 11 (0.9) | 0 | 0 | 1 (1.6) |  |
| **Aboriginal or Torres Strait islander** | 28 (2.2) | 1 (2.9) | 3 (2.8) | 0 | 0.36 (0.02-5.6) |
| **1<sup>st</sup> Quintile IRSAD** | 579 (45.3) | 14 (41.2) | 38 (35.8) | 31 (49.2) | 1.2 (0.72-1.9) |
| **Risk Category during acute admission (Purple or Blue)<sup>#</sup>** | 40 (3.2) | 1 (2.9) | 4 (3.8) | 3 (4.8) | 1.6 (0.49-4.6) |
| **Medical comorbidity** | 319 (25.0) | 10 (29.4) | 51 (48.1) | 28 (44.4) | 2.3 (1.4-3.7) |

Abbreviations: PCC, post-COVID condition; IRSAD, Index of relative socio-economic advantage and disadvantage.
\*Data reported as n (%) in sub-category unless otherwise specified.
^Relative risk of variable comparing cases flagged, reviewed and confirmed PCC with responders not flagged for concern.
<sup>#</sup>Children were assigned to blue (moderate), or purple (high) risk categories due to high number of risk factors and/or met clinical criteria (e.g. high fever, decreased O2 saturation).

## Discussion

In this study we assessed the medium-term outcome of acute COVID-19 in children and adolescents with laboratory confirmed SARS-CoV-2 infection in metropolitan Sydney following a discrete COVID outbreak with the Delta VoC in 2021. Several key findings have emerged. A great majority of respondents showed self-reported complete recovery at 12 weeks following infection. Amongst those who reported persistent symptoms, a wide spectrum was evident, and we conclude that Post COVID conditions (PCC) in children are both identifiable and heterogeneous. Those with features compatible with the UK consensus definition for Long COVID made up only a small subset of children and adolescents with persistent symptoms. Other subsets include those with a worsening of prior medical or developmental/behavioural co-morbidities, and those with emergent and persistent respiratory symptoms (albeit mild and without significant impact on daily function).

Reported prevalence estimates of PCC in children and adolescents have varied considerably with variability dependent on definitions used, location, presence or absence of controls groups.^10,11^ One systematic review of symptom prevalence in controlled follow-up studies (>4 weeks following COVID-19) showed significant risk differences (difference in symptom prevalence between cases of SARS-CoV-2 infection and controls without known SARS-CoV-2 infection) of 2-8% for individual symptoms including loss of smell, headaches, cognitive difficulties, sore throat and sore eyes; fatigue risk difference was 7% but not statistically significant.^22^ In the two included studies examining combinations of symptoms, the risk differences for 3 or more symptoms were 5% and 14%. Another, more recent, systematic review showed of 6 studies reported the prevalence difference was less than 4%.^11^ In these reviews, several studies reported symptom prevalence >4 weeks from infection, and only a minority in those only >12 weeks from infection. A large, national study from Denmark, not included in these prior reviews, reported a significantly higher prevalence of at least one symptom persisting more than 2 months in children and adolescents following confirmed SARS-CoV-2 infection when compared with controls. The difference was greatest in the youngest children; 0–3 years 12.8% difference, 4–11 years 4.4%, and 12–14 years 4.7%.^23^ The impact of vaccination on Long COVID prevalence and the effect newer variants have not been evaluated in children. We note emerging evidence that vaccination reduces prevalence, that is also potentially reduced following Omicron VoC infection.^24^

The accuracy of our prevalence estimates of PCC is limited by the response rate to our questionnaire and the absence of a non-infected control group in our study. However, our estimate for any persistent symptoms of concern of 6%, symptoms impacting on function of 3% and Long COVID (UK consensus definition compatible) of 1.2% are relatively low compared to other uncontrolled studies in the published literature and align well with those estimates from controlled studies.^10,22,23^ Further, our responder cohort showed broad similarity across measured demographic variables with our non-responder cohort. Where differences were evident in medical comorbidity and socioeconomic status, the current literature suggests these factors are likely to be associated with an increased risk of PCC.^22,25^ We therefore consider it likely that any bias introduced by these differences would result in an overestimation of PCC prevalence rather than underestimation.

There are few studies to help determine the risk factors for and overall severity of PCC in children and adolescents. Older age, female sex, and medical comorbidity have been associated with increased risk of persistent symptoms in two systematic reviews. ^22,25^ Our study adds support to older age and medical co-morbidity being risk factors for PCC, and female sex was over-represented in children clinically considered to have Long COVID.

Few studies have quantified impact of persistent symptoms post COVID in children on functioning or measures of HRQoL. In a Danish study^23^, differences in health-related quality of life (PedsQL) scores were seen in younger children with worse physical symptom scores in confirmed SARS-CoV-2 infected children than in uninfected controls. In a population based Norwegian study, children (aged <16 years) following SARS-CoV-2 infection showed a relative increased use of primary health services but not specialist outpatient or hospital services compared with uninfected controls for 3 months.^26^ The effect was largest and lasted longest (to 6 months) in children aged 1-5 years. In our study, the relative difference between children reporting persistent symptoms and those in whom it was reported to impact on function or result in seeking additional health assessment adds support to the view that a considerable proportion of PCC in children are mild. In the children clinically reviewed, a majority had already recovered or reported single-organ symptoms which were not impacting on daily activities including return to school. The data from Magnusson et al.^26^ also suggest a natural history of PCC of recovery over a 6 month period, although the longitudinal course of PCC needs further study. Recently published results from a longitudinal analysis of symptom prevalence from the UK CLoCK study showed that a majority of measured symptoms reduced in prevalence between 3 and 6 months post COVID suggesting a trajectory to recovery of PCC is likely across this time period.^27^

Our data suggest that Long COVID as a debilitating, multi-system illness, that is experienced by only a minority of children with persistent symptoms 12 weeks following acute COVID-19, determined by a survey/questionnaire approach. We identified a relatively greater number of children with single symptom, mainly respiratory, upper and lower tract, persistent problems including loss of taste/smell. Addressing the needs of these children is also of importance beyond the consideration of Long COVID specifically. We have also shown evidence of COVID-19 exacerbating the symptoms of pre-existing medical, developmental-behavioural and mental health conditions in children. We cannot determine whether these effects are direct or indirect, although the impression of the clinicians participating in this work was that it was more likely to have arisen indirectly from the disruption of the COVID-19 pandemic more broadly or the disruptions to the child/family from the isolation requirements of the acute illness. This additional disease burden may be attributed directly to COVID-19 from the perspective of patients/families and the wider community.

Although children on clinical review who met the clinical definition of Long COVID only represent a small proportion of our responder cohort, this proportion potentially constitutes a large number of children where population wide infection occurs. Serosurvey data from Australia showed that the only ∼2% of the child population showed serological evidence of infection following the Delta VoC wave (Koirala et al. MJA 2022); however a recent survey performed after 7 months of Omicron VoC transmission showed 60-80% of children had been infected (see: 2022 serosurvey summary report at https://ncirs.org.au/reports). Although we do not know whether PCC following Omicron VoCs are similar in frequency to Delta in our setting, the absolute number of infections indicates a need for scalable person-centred interventions is now more pressing. Health services need to develop pathways to support screening of children for non-recovery and provide better guidance for primary care services to support detection, initial case management and improved pathways to escalation of care and thereby provided tailored treatment strategies to deliver the right care at the right time. At present, Australia lacks a comprehensive approach to collect data on PCC across the lifespan. Without such data evaluation of the prevalence and severity of PCC with current and future variants in the Australian context will be hampered, as well as a more comprehensive evaluation of long-term outcomes and the burden on the health system. Also at present, the management of PCC, including Long COVID, is empiric by analogy with other causes of post-viral fatigue states. Without better case identification and tracking in Australia, implementation of interventional studies will not be possible.

Our study has several strengths: all cases of SARS-CoV-2 infection were laboratory confirmed by PCR testing in a context of very high-test frequency at a population level. Furthermore, prior to the Delta VoC outbreak infection rates in children generally in New South Wales had been extremely low with COVID-19 well controlled, potentially allowing for better assessment of the direct effect of COVID-19 itself on post-COVID symptoms, rather than indirect pandemic effects. The follow-up questionnaire included explicit evaluation of responses relative to the child’s acute SARS-CoV-2 illness and all cases had been managed during their acute infection through a structured ambulatory care model.^15^ Our study sought responses from all aged children; the inclusion of children aged <5 years has been identified as a gap in the already limited literature on PCC in children.^28,29^ In addition to a standardised symptom list, our questionnaire collected data on unmet healthcare need, HRQoL and school attendance. An additional strength was the clinician review and validation of responses amongst >80% of cases who were flagged for symptoms of concern, and the application of the UK consensus case definition for long COVID in children.

Despite these strengths, our study has several limitations. Firstly, we did not have a control group of either SARS-CoV-2 test-negative children or children with another respiratory virus. This is a well-recognised issue in the field, but again we point to the relatively low prevalence of persistent symptoms identified in our responder cohort that aligns with risk differences reported in controlled studies. Secondly, our survey response rate was <20%. Again, this is a recognised weakness of other questionnaire-based studies in the field - the 3-month response rate in the UK CLoCK study was 13.4% -, but we point to the broad similarity of our responder cohort with the measured characteristics of the non-responder cohort. Thirdly, amongst respondents, beyond the initial screening questions, the full questionnaire was completion was variable. Fourthly, the questionnaire was only available in English that may have impacted on the capacity for children and families from culturally and linguistically diverse backgrounds to respond. Lastly, the clinical review was undertaken via telehealth in almost all cases and clinical categorisation was pragmatic and post-hoc.

In conclusion, we have described a low estimated prevalence, but broad spectrum and severity of post-COVID conditions at >12 weeks following predominantly mild SARS-CoV-2 infection in children aged <16 years.

## Data Availability

All data produced in the present study are available upon reasonable request to the authors

## Acknowledgements

We acknowledge the many clinicians (nursing, medical and allied health) who contributed to the acute care of children with SARS-CoV-2 infection under the auspices of the SCHN virtualKIDS COVID□19 outpatient response team.

## Supplementary Materials

**Supplementary Figure 1:**
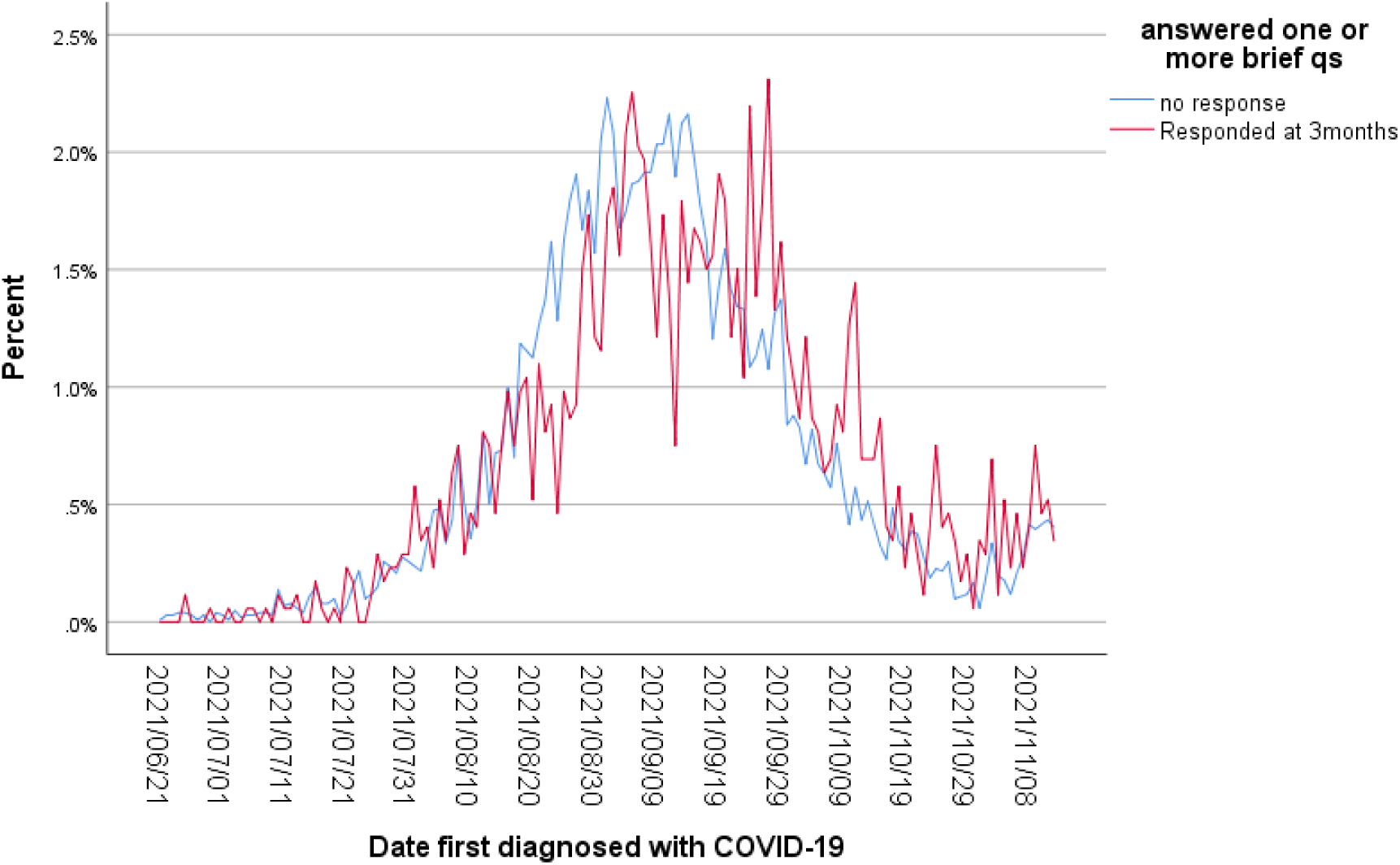
Daily percentage of responder and non-responder cohorts to follow-up questionnaire by date of SARS-CoV-2 infection week across the 2021 Delta VoC outbreak in Metropolitan Sydney.

**Supplementary Table 1:**
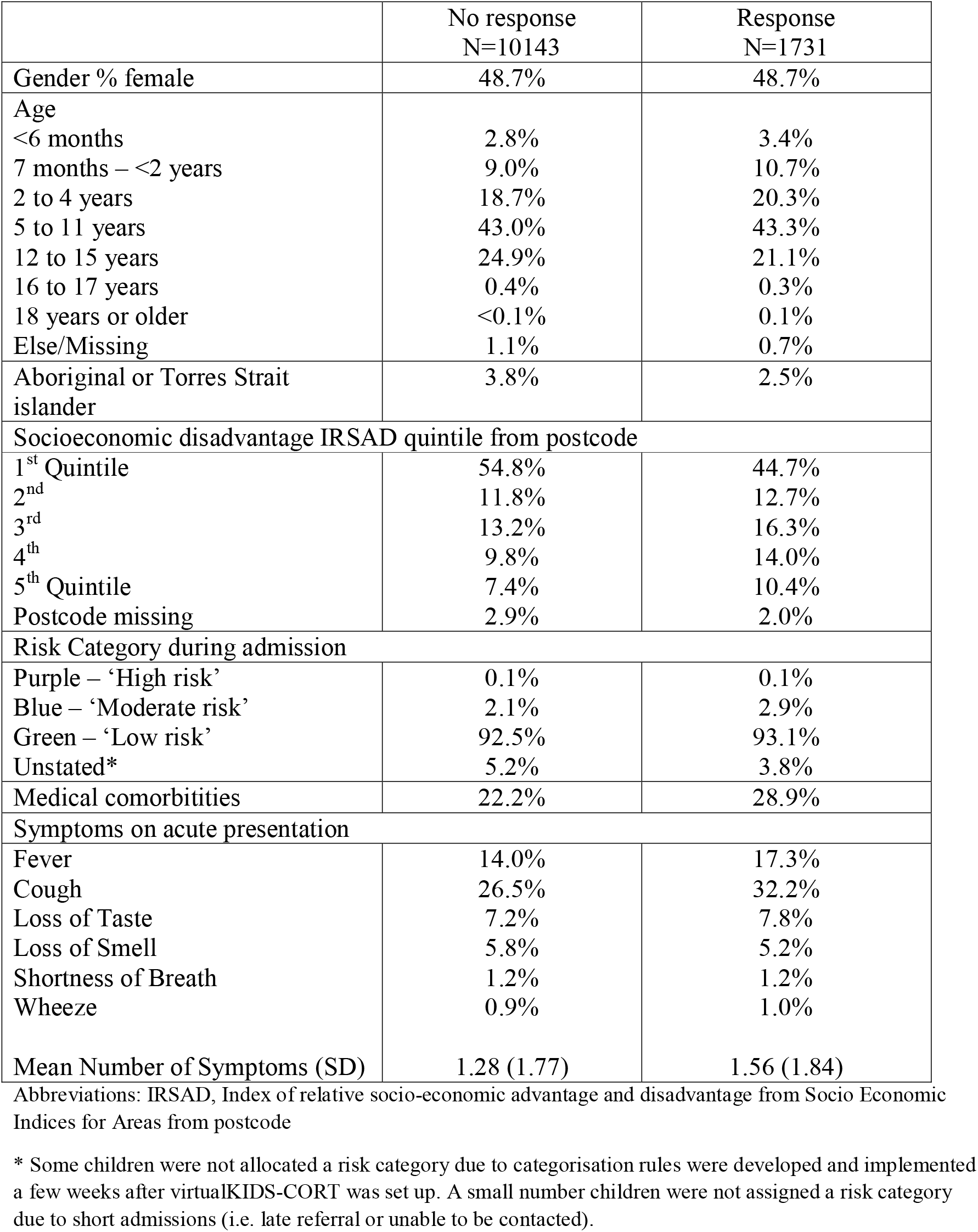
Demographic, acute severity and acute symptom profile of responders and non-responders to follow-up questionnaire.

|  | No response<br>N=10143 | Response<br>N=1731 |
| --- | --- | --- |
| Gender % female | 48.7% | 48.7% |
| Age |  |  |
| <6 months | 2.8% | 3.4% |
| 7 months – <2 years | 9.0% | 10.7% |
| 2 to 4 years | 18.7% | 20.3% |
| 5 to 11 years | 43.0% | 43.3% |
| 12 to 15 years | 24.9% | 21.1% |
| 16 to 17 years | 0.4% | 0.3% |
| 18 years or older | <0.1% | 0.1% |
| Else/Missing | 1.1% | 0.7% |
| Aboriginal or Torres Strait islander | 3.8% | 2.5% |
| Socioeconomic disadvantage IRSAD quintile from postcode |  |  |
| 1<sup>st</sup> Quintile | 54.8% | 44.7% |
| 2<sup>nd</sup> | 11.8% | 12.7% |
| 3<sup>rd</sup> | 13.2% | 16.3% |
| 4<sup>th</sup> | 9.8% | 14.0% |
| 5<sup>th</sup> Quintile | 7.4% | 10.4% |
| Postcode missing | 2.9% | 2.0% |
| Risk Category during admission |  |  |
| Purple – ‘High risk’ | 0.1% | 0.1% |
| Blue – ‘Moderate risk’ | 2.1% | 2.9% |
| Green – ‘Low risk’ | 92.5% | 93.1% |
| Unstated* | 5.2% | 3.8% |
| Medical comorbidities | 22.2% | 28.9% |
| Symptoms on acute presentation |  |  |
| Fever | 14.0% | 17.3% |
| Cough | 26.5% | 32.2% |
| Loss of Taste | 7.2% | 7.8% |
| Loss of Smell | 5.8% | 5.2% |
| Shortness of Breath | 1.2% | 1.2% |
| Wheeze | 0.9% | 1.0% |
| Mean Number of Symptoms (SD) | 1.28 (1.77) | 1.56 (1.84) |

Abbreviations: IRSAD, Index of relative socio-economic advantage and disadvantage from Socio Economic Indices for Areas from postcode
\* Some children were not allocated a risk category due to categorisation rules were developed and implemented a few weeks after virtualKIDS-CORT was set up. A small number children were not assigned a risk category due to short admissions (i.e. late referral or unable to be contacted).

**Supplementary Table. 2.**
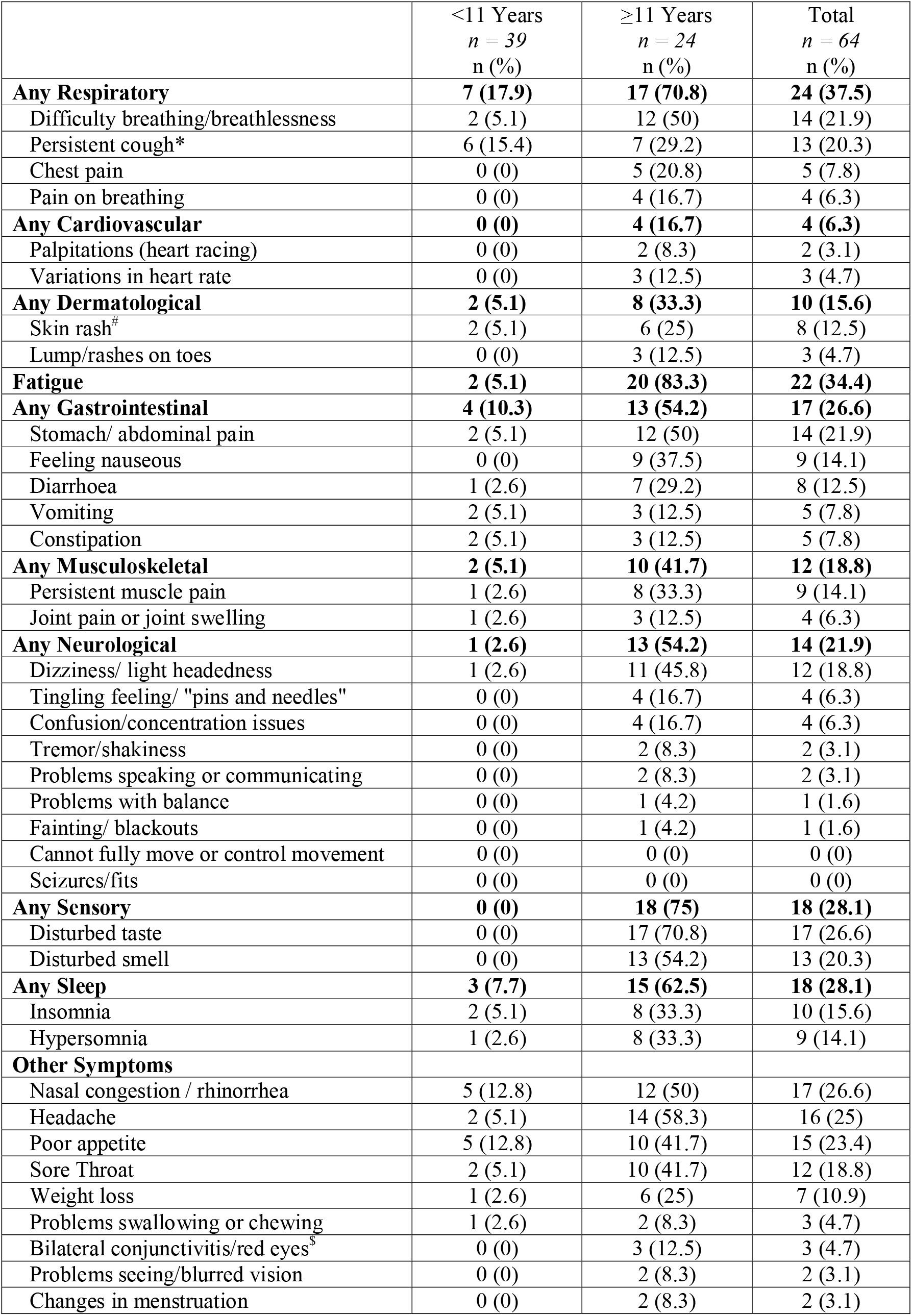
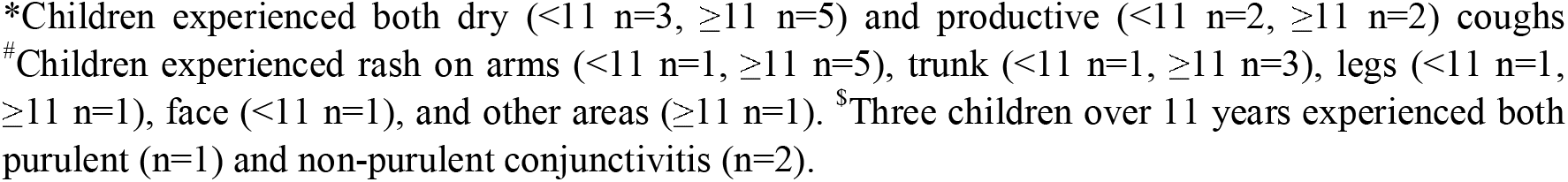
Self-reported persistent symptoms using ISARIC symptom questionnaire symptom list (n=64).

|  | <11 Years<br>*n* = 39<br>*n* (%) | ≥11 Years<br>*n* = 24<br>*n* (%) | Total<br>*n* = 64<br>*n* (%) |
| --- | --- | --- | --- |
| **Any Respiratory** | **7 (17.9)** | **17 (70.8)** | **24 (37.5)** |
| Difficulty breathing/breathlessness | 2 (5.1) | 12 (50) | 14 (21.9) |
| Persistent cough* | 6 (15.4) | 7 (29.2) | 13 (20.3) |
| Chest pain | 0 (0) | 5 (20.8) | 5 (7.8) |
| Pain on breathing | 0 (0) | 4 (16.7) | 4 (6.3) |
| **Any Cardiovascular** | **0 (0)** | **4 (16.7)** | **4 (6.3)** |
| Palpitations (heart racing) | 0 (0) | 2 (8.3) | 2 (3.1) |
| Variations in heart rate | 0 (0) | 3 (12.5) | 3 (4.7) |
| **Any Dermatological** | **2 (5.1)** | **8 (33.3)** | **10 (15.6)** |
| Skin rash<sup>#</sup> | 2 (5.1) | 6 (25) | 8 (12.5) |
| Lump/rashes on toes | 0 (0) | 3 (12.5) | 3 (4.7) |
| **Fatigue** | **2 (5.1)** | **20 (83.3)** | **22 (34.4)** |
| **Any Gastrointestinal** | **4 (10.3)** | **13 (54.2)** | **17 (26.6)** |
| Stomach/ abdominal pain | 2 (5.1) | 12 (50) | 14 (21.9) |
| Feeling nauseous | 0 (0) | 9 (37.5) | 9 (14.1) |
| Diarrhoea | 1 (2.6) | 7 (29.2) | 8 (12.5) |
| Vomiting | 2 (5.1) | 3 (12.5) | 5 (7.8) |
| Constipation | 2 (5.1) | 3 (12.5) | 5 (7.8) |
| **Any Musculoskeletal** | **2 (5.1)** | **10 (41.7)** | **12 (18.8)** |
| Persistent muscle pain | 1 (2.6) | 8 (33.3) | 9 (14.1) |
| Joint pain or joint swelling | 1 (2.6) | 3 (12.5) | 4 (6.3) |
| **Any Neurological** | **1 (2.6)** | **13 (54.2)** | **14 (21.9)** |
| Dizziness/ light headedness | 1 (2.6) | 11 (45.8) | 12 (18.8) |
| Tingling feeling/ "pins and needles" | 0 (0) | 4 (16.7) | 4 (6.3) |
| Confusion/concentration issues | 0 (0) | 4 (16.7) | 4 (6.3) |
| Tremor/shakiness | 0 (0) | 2 (8.3) | 2 (3.1) |
| Problems speaking or communicating | 0 (0) | 2 (8.3) | 2 (3.1) |
| Problems with balance | 0 (0) | 1 (4.2) | 1 (1.6) |
| Fainting/ blackouts | 0 (0) | 1 (4.2) | 1 (1.6) |
| Cannot fully move or control movement | 0 (0) | 0 (0) | 0 (0) |
| Seizures/fits | 0 (0) | 0 (0) | 0 (0) |
| **Any Sensory** | **0 (0)** | **18 (75)** | **18 (28.1)** |
| Disturbed taste | 0 (0) | 17 (70.8) | 17 (26.6) |
| Disturbed smell | 0 (0) | 13 (54.2) | 13 (20.3) |
| **Any Sleep** | **3 (7.7)** | **15 (62.5)** | **18 (28.1)** |
| Insomnia | 2 (5.1) | 8 (33.3) | 10 (15.6) |
| Hypersomnia | 1 (2.6) | 8 (33.3) | 9 (14.1) |
| **Other Symptoms** |  |  |  |
| Nasal congestion / rhinorrhea | 5 (12.8) | 12 (50) | 17 (26.6) |
| Headache | 2 (5.1) | 14 (58.3) | 16 (25) |
| Poor appetite | 5 (12.8) | 10 (41.7) | 15 (23.4) |
| Sore Throat | 2 (5.1) | 10 (41.7) | 12 (18.8) |
| Weight loss | 1 (2.6) | 6 (25) | 7 (10.9) |
| Problems swallowing or chewing | 1 (2.6) | 2 (8.3) | 3 (4.7) |
| Bilateral conjunctivitis/red eyes<sup>s</sup> | 0 (0) | 3 (12.5) | 3 (4.7) |
| Problems seeing/blurred vision | 0 (0) | 2 (8.3) | 2 (3.1) |
| Changes in menstruation | 0 (0) | 2 (8.3) | 2 (3.1) |

\*Children experienced both dry ( $<11$ $n=3$ , $\geq 11$ $n=5$ ) and productive ( $<11$ $n=2$ , $\geq 11$ $n=2$ ) coughs #Children experienced rash on arms ( $<11$ $n=1$ , $\geq 11$ $n=5$ ), trunk ( $<11$ $n=1$ , $\geq 11$ $n=3$ ), legs ( $<11$ $n=1$ , $\geq 11$ $n=1$ ), face ( $<11$ $n=1$ ), and other areas ( $\geq 11$ $n=1$ ). \$Three children over 11 years experienced both purulent ( $n=1$ ) and non-purulent conjunctivitis ( $n=2$ ).
\*Children experienced both dry ( $<11$ $n=3$ , $\geq 11$ $n=5$ ) and productive ( $<11$ $n=2$ , $\geq 11$ $n=2$ ) coughs

**Supplementary Figure 2:**
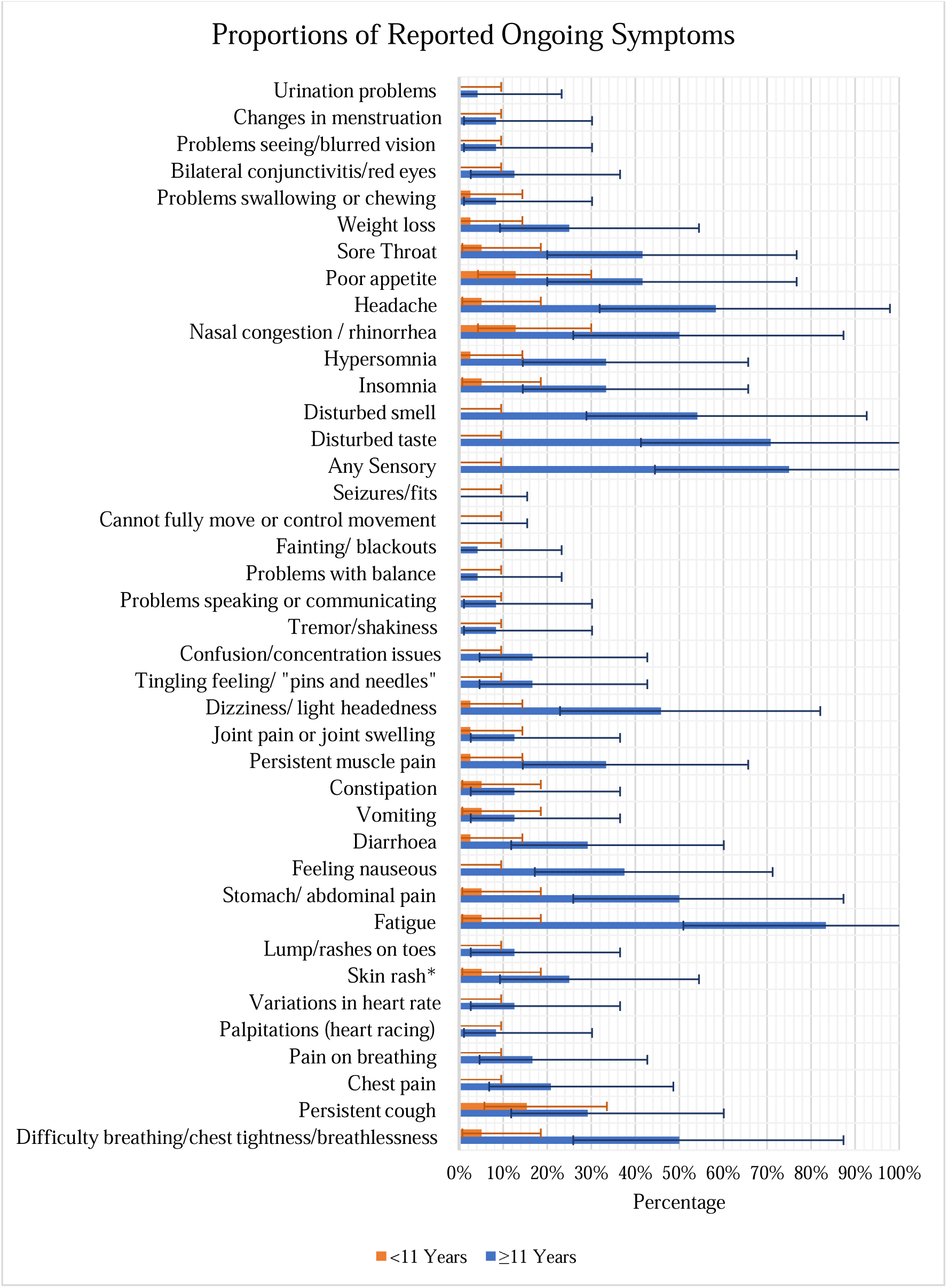
Bar graph of self-reported persistent symptoms (percentage and 95% confidence intervals) using ISARIC symptom questionnaire symptom list stratified by age; younger children aged <11yrs, older children aged ≥11yrs (n=64).

**Supplementary Table. 3.** Selected additional results from questionnaire responses.

| Key question (response result) | n/N | % (95% CI ) |
| --- | --- | --- |
| During the last 30 days, about how often did [your child] feel that everything was an effort? (>='most of the time') | 19/409 | 4.7 (2.8-7.2) |
| Has there been a change in [your child's] usual mood since their COVID-19 illness? (YES) | 146/1569 | 9.3 (7.9-10.9) |
| Has [your child] been able to return to their normal activities without problems since their COVID-19 illness? (NO) | 73/1569 | 4.7 (3.7-5.8) |
| In the past 30 days have you asked for help from a health professional about [your child's] COVID-19 illness? (YES) | 58/1559 | 3.7 (2.8-4.8) |
| Has [your child] visited a doctor/health centre because of COVID-19 health consequences? (YES) | 79/1559 | 5.1 (4.0-6.3) |
| Has [your child] had any symptoms in the past 7 days which were not present before the start of their COVID-19 illness? (YES) | 65/1547 | 4.2 (3.3-5.3) |
| Health score TODAY? (0-100 scale) | N=655<br>Mean 87.8<br>Median 95 | (SD 17.6)<br>(IQR 82;100) |
| Health score BEFORE COVID-19 (0-100 scale) | N=655<br>Mean 90.7<br>Median 97 | (SD 14.3)<br>(IQR 89;100) |
| K6 total score | N=410<br>Mean 8.3<br>Median 6<br>>13 = 10% | (SD 3.7)<br>(IQR 6;9) |

